# A Video Intervention to Improve Patient Understanding of Tumor Genomic Testing in Patients with Cancer

**DOI:** 10.1101/2023.12.05.23299443

**Authors:** Deloris Veney, Lai Wei, Amanda E. Toland, Carolyn J. Presley, Heather Hampel, Tasleem J. Padamsee, Clara N. Lee, William J. Irvin, Michael Bishop, James Kim, Shelly R. Hovick, Leigha Senter, Daniel G. Stover

## Abstract

**Background:** Tumor genomic testing (TGT) has become standard-of-care for most patients with advanced/metastatic cancer. Despite established guidelines, patient education prior to TGT is variable or frequently omitted. The purpose of this study was to evaluate the impact of a concise (3-4 minute) video for patient education prior to TGT.

**Methods:** Based on a quality improvement cycle, an animated video was created to be applicable to any cancer type, incorporating culturally diverse images, available in English and Spanish. Patients undergoing standard-of care TGT were enrolled at a tertiary academic institution and completed validated survey instruments immediately prior to video viewing (T1) and immediately post-viewing (T2). Instruments included: 1) 10-question objective genomic knowledge/understanding; 2) 10-question video message-specific knowledge/recall; 3) 11-question Trust in Physician/Provider; 4) attitudes regarding TGT. The primary objective was change in outcomes from before to after the video was assessed with Wilcoxon signed rank test.

**Results:** From April 2022 to May 2023, a total of 150 participants were enrolled (MBC n=53, LC n=38, OC n=59). For the primary endpoint, there was a significant increase in video message-specific knowledge (median 10 point increase; p<0.0001) with no significant change in genomic knowledge/understanding (p=0.89) or Trust in Physician/Provider (p=0.59). Results for five questions significantly improved, including the likelihood of TGT impact on treatment decision, incidental germline findings, and cost of testing. Improvement in video message-specific knowledge was consistent across demographic groups, including age, income, and education. Individuals with less educational attainment had had greater improvement from before to after video viewing.

**Conclusions:** A concise, 3-4 minute, broadly applicable video incorporating culturally diverse images administered prior to TGT significantly improved video message-specific knowledge across all demographic groups. This resource is publicly available at http://www.tumor-testing.com, with a goal to efficiently educate and empower patients regarding TGT while addressing guidelines within the flow of clinical practice.

**Clinical Trial Registration:** ClinicalTrials.gov NCT05215769

## Introduction

Somatic next-generation sequencing, also known as tumor genomic testing (TGT) has become increasingly adopted as part of standard cancer care for many cancers.^1^ From 2017-2021, there was an estimated 3-fold increase (9% to 30%) in the proportion of tumors for which there was a TGT-identified mutation with a disease-matched, standard-care, FDA-approved therapy.^2^ As a result of three molecular alterations each with tumor-agnostic FDA approved therapies (*NTRK* fusion – larotrectinib; deficient mismatch repair – pembrolizumab; high tumor mutational burden - pembrolizumab), TGT is now viewed as a necessity by most oncologists in nearly all patients with metastatic cancer.^2^ As a result, its use has become widespread in the metastatic setting and has increased in some early stage settings.

International guidelines all recommend that clinicians report incidental germline findings and likely germline findings (e.g., from tumor-normal TGT or pathogenic variants in germline-relevant genes at high allele frequencies) to their patients.^3-10^ The American Society of Clinical Oncology (ASCO) Policy Statement^3^ notes: “1) Oncology providers should communicate the potential for incidental/secondary germline information…before conducting somatic mutation profiling and should review potential benefits, limitations, and risks before testing; 2) Providers should carefully ascertain patient preferences regarding the receipt of germline information.” Despite these unified guidelines, evidence suggests that provider-patient discussions around TGT are inconsistent,^11,12^ which is complicated further by limited genetics/genomics literacy among patients,^13^ particularly those who have lower income and/or those who are medically underserved.^14^ This raises important ethical challenges including uncertainty of results, incidental germline findings, and disparities around TGT options and access.^11,15,16,17^

Taken together, this evidence supports the need for consistent and improved communication between providers and patients about TGT. To address this need, we previously conducted a quality improvement (QI) initiative focused on patient education prior to TGT using a Plan-Do-Study-Act (PDSA) approach.^18,19^ Within that PDSA cycle, published guidelines related to pre-TGT provider-patient education were reviewed; a provider QI survey highlighted inconsistency in pre-TGT discussion practice across providers; and patient focus groups and interviews revealed important themes and opportunities. Themes and opportunities were incorporated into a patient-navigated, concise 3-4 minute animated video for pre-TGT education with content addressing 14/17 (82%) of key points described in the ASCO and ACMG guidelines.^3,6,19^ The video is based on adult learning and communication theory and includes characters of varied races, ethnicities, and genders so that the images presented are relatable to patients of varied identities.

We report the primary outcome analysis of a prospective study evaluating the impact of this concise pre-TGT educational video intervention on video message-specific knowledge, general genomic knowledge/understanding, and trust in physician/provider.

## Methods

### Patient Eligibility and Recruitment

The study protocol was approved by the Ohio State University Institutional Review Board (OSU#2021C0209). Participants were eligible for inclusion if they were age eighteen or older at the time of study entry with biopsy-confirmed cancer, spoke English or Spanish, and planned to undergo TGT. TGT could be from tumor tissue or blood-based. Any commercial vendor of TGT or Ohio State University Molecular Pathology lab was acceptable. Eligible patients receiving care in cancer care clinics at The Ohio State University’s James Cance Center facilities were identified by research staff through TGT order placement, screening, or provider referral. A total of 156 participants were consented to achieve the prespecified 150 enrolled subjects completing all required surveys to be eligible for inclusion in the primary analysis (CONSORT diagram **Supp Fig 1A**) from March 2022 through May 2023 (**Supp Fig 1B**).

### Survey Instruments

<u>Video message–specific knowledge (VMSK)</u>: Message-specific knowledge was measured 10 true/false statements that addressed key knowledge domains in the video intervention, with a final score reported as number correct multiplied by 100. Examples of statements include “My doctor might recommend that family members undergo genetic testing based on the result of this tumor test”, “The result of my tumor test might not change my treatment.” <u>Genomic Knowledge/Understanding</u>: Objective knowledge of genes/genetics was measured with 10 true/false statements based on a published genetic knowledge instrument, with a final score reported as number correct multiplied by 100.^20^ Examples of these statements include “It is possible to see a gene with the naked eye”, and “A person’s race and ethnicity can affect how likely they are to get a disease”. <u>Trust in Physician/Provider (TIPP)</u>: The 11-item Trust in Physician/Provider Survey^21^ uses a 5-point Likert scalet. TPS scores range from 11 to 55 with higher scores indicating greater trust in provider.^21^

### Study Procedures

All study procedures, including informed consent, survey instruments, and video intervention were completed through a single REDCap survey via tablet. Both English and Spanish versions of the survey instruments were provided as applicable. Participants were enrolled across three cohorts, with no mandated minimum or maximum number of patients per cohort: Cohort 1. Metastatic breast cancer (MBC); Cohort 2. Lung cancer (LC); Cohort 3. Other cancer of any type (OC). The video viewed by each cohort differed by the modular adaptation applied to the video: OC cohort participants viewed a 3.05-minute video; MBC cohort participants viewed the same video with the addition of a sixteen second clip indicating that at most four out of 10 patients with MBC receive a tumor genomic test result that determines their treatment; LC cohort participants also had an additional sixteen seconds of video content indicating that three out of 10 patients with LC have a tumor genomic test result that determines their treatment. Survey instruments were completed at timepoint 1 (T1), immediately prior to video viewing, including: demographics, genomic knowledge/understanding, video message-specific knowledge, Trust in Physician/Provider, attitudes around genomic testing, and intentions regarding TGT. None of the knowledge-related questions were cohort-specific. With the exception of demographics, all instruments were repeated at timepoint 2 (T2), following video viewing and prior to discussion with provider of participant’s own results. Additionally, at T2, participants completed an opinion assessment of the video itself. Survey questions are provided in **Supplementary File 1**.

### Statistical Analyses

Only patients who completed all T1 and T2 surveys were included for analyses. The primary objective was to assess change in video message-specific knowledge from pre- to post-exposure to the TGT educational video. Secondary endpoints included: 1) change in video message-specific knowledge within each cohort (MBC, LC, or OC); 2) change in genomic knowledge/understanding in the overall study population and within each cohort; 3) change in Trust in Physician/Provider as a single score in the overall study population and within each cohort. Based on preliminary data, with a cohort of 150 patients, there would be 90% power to detect an effect size of 0.66 in change of recall accuracy from pre-(immediately prior) to post-(immediately after) video intervention, using a two-sided Wilcoxon signed-rank test with alpha of 0.05. All secondary outcomes were summarized using descriptive statistics and compared pre-/post-video using Wilcoxon signed-rank test, which indicates…. Evaluation of change in proportion of individuals answering specific questions correctly within video message-specific knowledge was assessed using McNemar’s test for the whole population and within each cohort. The associations of video message-specific knowledge, genomic knowledge/understanding, and Trust in Physician/Provider with age were explored with Spearman correlations and the associations of video message-specific knowledge, genomic knowledge/understanding, and Trust in Physician/Provider with other categorical demographics were explored with Kruskal-Wallis test; given the exploratory nature of these analyses no multiple test correction was used and nominal p-values were reported.

## Results

### Study Participants

150 participants completed survey instruments at both T1 and T2 and were considered evaluable for the primary endpoint. Participant characteristics at baseline by cohort are provided in **Table 1**. In the total cohort, participant age ranged from 18 to 93 years at study entry. Most participants were female (94/150; 63%), of White race (132/150, 88%), and were married or in a domestic partnership (102/150, 62%). Education was relatively evenly distributed among high school or less, some college or technical school, and college or graduate degree, while income was similarly relatively evenly distributed from self-reported annual income less than $25,000 to greater than $75,000 (**Table 1**). Among participants, eight distinct cancer types were represented including: breast cancer, lung cancer, and gastrointestinal cancer most common (**Supp Fig 1C**). Genomic testing vendors included Tempus (n=46), Foundation Medicine (n=41), Guardant (n=20), Caris Life Sciences (n=13), Natera (n=12), and Ohio State University Molecular Pathology (n=18) (**Supp Fig 1D**).

**Table 1.**
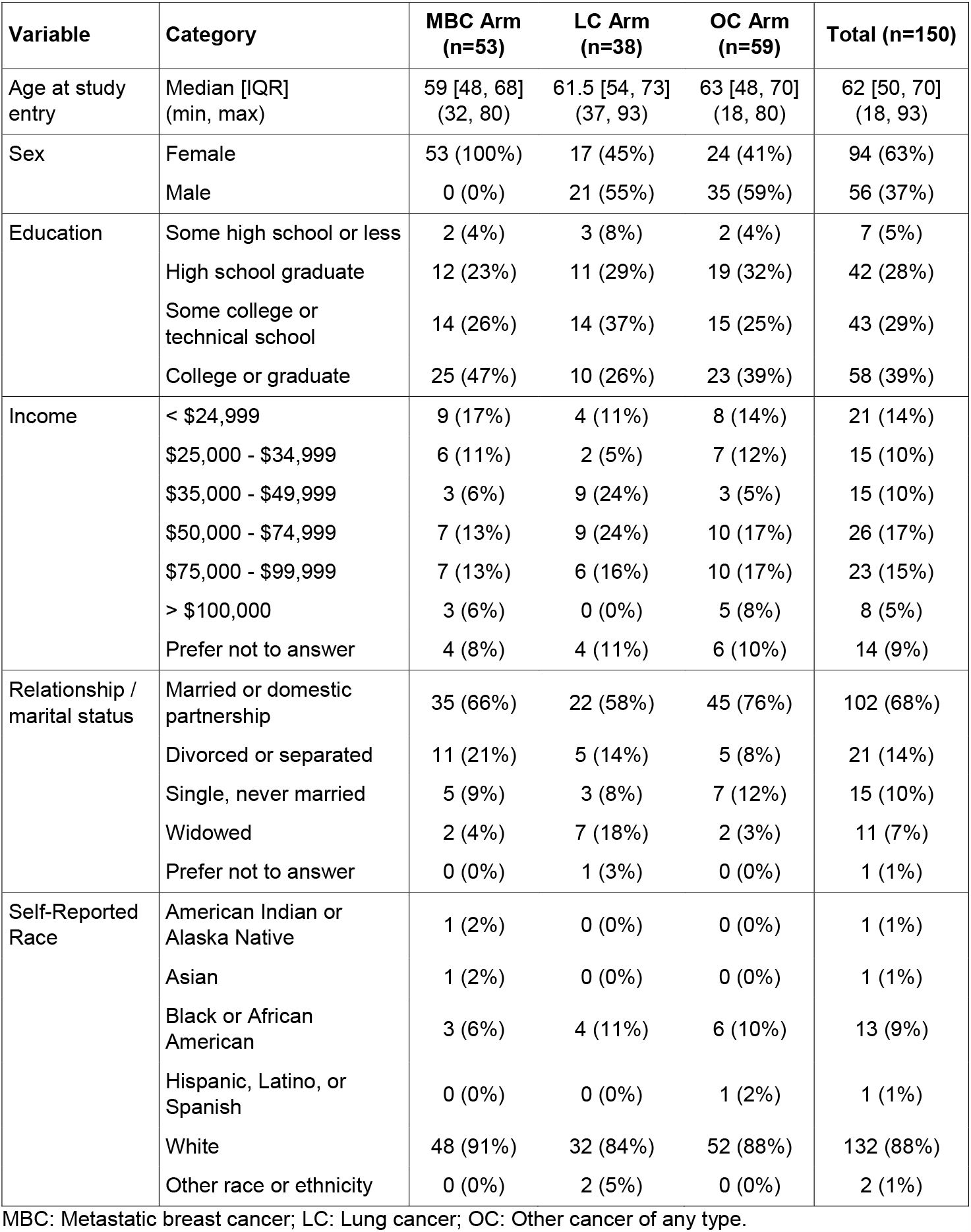
Participant Characteristics at Baseline.

| Variable | Category | MBC Arm<br>(n=53) | LC Arm<br>(n=38) | OC Arm<br>(n=59) | Total (n=150) |
| --- | --- | --- | --- | --- | --- |
| Age at study entry | Median [IQR]<br>(min, max) | 59 [48, 68]<br>(32, 80) | 61.5 [54, 73]<br>(37, 93) | 63 [48, 70]<br>(18, 80) | 62 [50, 70]<br>(18, 93) |
| Sex | Female | 53 (100%) | 17 (45%) | 24 (41%) | 94 (63%) |
|  | Male | 0 (0%) | 21 (55%) | 35 (59%) | 56 (37%) |
| Education | Some high school or less | 2 (4%) | 3 (8%) | 2 (4%) | 7 (5%) |
|  | High school graduate | 12 (23%) | 11 (29%) | 19 (32%) | 42 (28%) |
|  | Some college or technical school | 14 (26%) | 14 (37%) | 15 (25%) | 43 (29%) |
|  | College or graduate | 25 (47%) | 10 (26%) | 23 (39%) | 58 (39%) |
| Income | < \$24,999 | 9 (17%) | 4 (11%) | 8 (14%) | 21 (14%) |
| | \$25,000 - \$34,999 | 6 (11%) | 2 (5%) | 7 (12%) | 15 (10%) |
| | \$35,000 - \$49,999 | 3 (6%) | 9 (24%) | 3 (5%) | 15 (10%) |
| | \$50,000 - \$74,999 | 7 (13%) | 9 (24%) | 10 (17%) | 26 (17%) |
| | \$75,000 - \$99,999 | 7 (13%) | 6 (16%) | 10 (17%) | 23 (15%) |
| | > \$100,000 | 3 (6%) | 0 (0%) | 5 (8%) | 8 (5%) |
|  | Prefer not to answer | 4 (8%) | 4 (11%) | 6 (10%) | 14 (9%) |
| Relationship / marital status | Married or domestic partnership | 35 (66%) | 22 (58%) | 45 (76%) | 102 (68%) |
|  | Divorced or separated | 11 (21%) | 5 (14%) | 5 (8%) | 21 (14%) |
|  | Single, never married | 5 (9%) | 3 (8%) | 7 (12%) | 15 (10%) |
|  | Widowed | 2 (4%) | 7 (18%) | 2 (3%) | 11 (7%) |
|  | Prefer not to answer | 0 (0%) | 1 (3%) | 0 (0%) | 1 (1%) |
| Self-Reported Race | American Indian or Alaska Native | 1 (2%) | 0 (0%) | 0 (0%) | 1 (1%) |
|  | Asian | 1 (2%) | 0 (0%) | 0 (0%) | 1 (1%) |
|  | Black or African American | 3 (6%) | 4 (11%) | 6 (10%) | 13 (9%) |
|  | Hispanic, Latino, or Spanish | 0 (0%) | 0 (0%) | 1 (2%) | 1 (1%) |
|  | White | 48 (91%) | 32 (84%) | 52 (88%) | 132 (88%) |
|  | Other race or ethnicity | 0 (0%) | 2 (5%) | 0 (0%) | 2 (1%) |

MBC: Metastatic breast cancer; LC: Lung cancer; OC: Other cancer of any type.

*Video Message-Specific Knowledge, Genomic Knowledge and Understanding, and Trust in Physician/Provider* For the primary endpoint, there was a significant increase in video message-specific knowledge score (sum of correct true-false questions multiplied by 100) from T1 to T2 (median increase (interquartile range/IQR: 10 (0,10) Wilcoxon signed rank p<0.0001) (**Fig 1A**). Concurrently, genomic knowledge/understanding score did not significantly change (Wilcoxon signed rank p=0.89 with median increase (IQR): 0 (-10,10) ; **Fig 1B**) and Trust in Physician/Provider score did not significantly change (Wilcoxon signed rank p=0.59 with median increase (IQR): 0 (-1, 1) ; **Fig 1C**). There were significant increases in video message-specific knowledge within each cohort (Wilcoxon signed rank p<0.0001 MBC; p<0.0001 LC; p<0.0001 OC; **Fig 2A**) with no significant change within any cohort for genomic knowledge/understanding (**Fig 2B**; all p>0.05) or Trust in Physician/Provider (**Fig 2B**; all p>0.05) (numerical data for Fig 2 provided as **Supp Table 1**).

**Figure 1.**
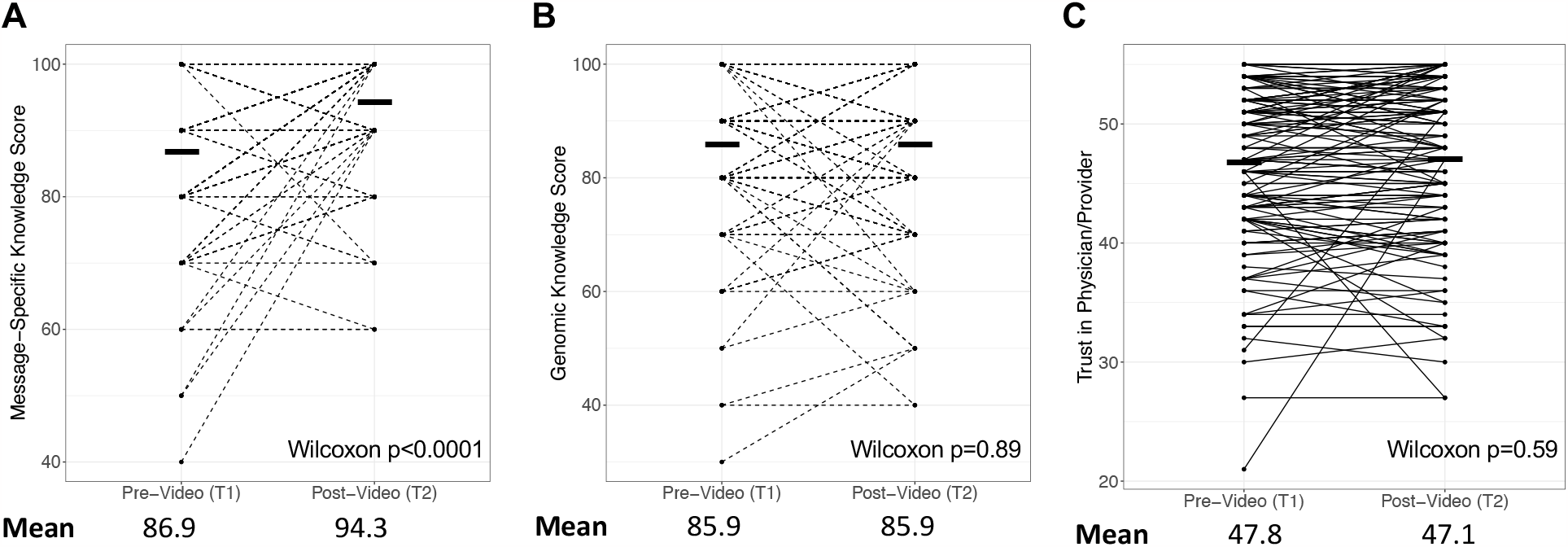
Change in Knowledge and Trust Metrics Pre- to Post-Video. Participants completed survey assessments pre-video viewing (T1) and post-video viewing (T2) of the 3-4 minute tumor genomic testing educational video intervention. Survey assessments included: 10-question video message-specific knowledge with score reported as number correct multiplied by 100 (VMSK; **A**), 10-question general genomic knowledge and understanding with score reported as number correct multiplied by 100 (GKU; **B**), and 11-question trust in physician/provider (TIPP; **C**). Paired scores for each participant are presented with mean and Wilcoxon signed rank. Change in video message-specific knowledge from T1 to T2 was the primary endpoint of the study

**Figure 2.**
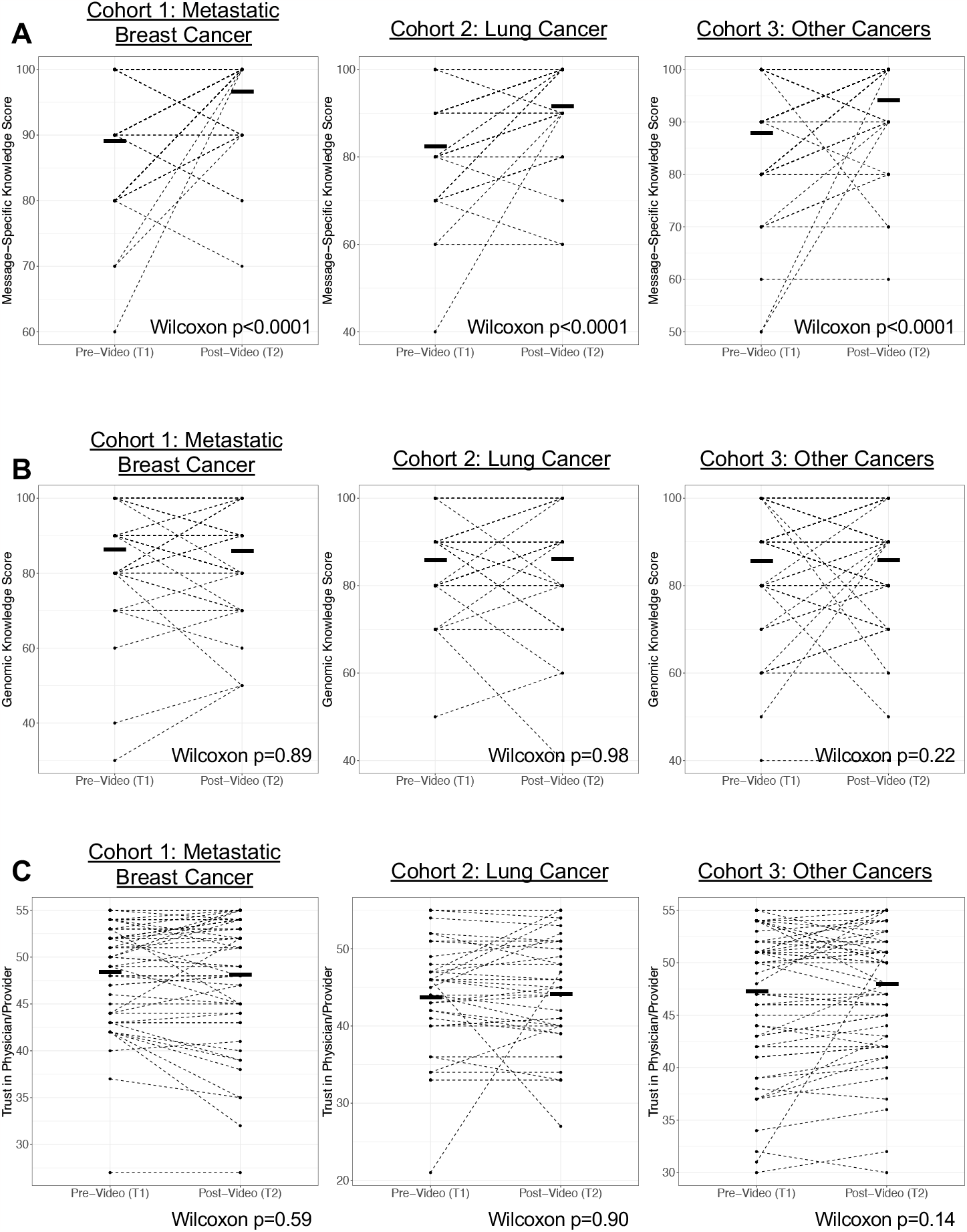
Within-Cohort Change in Knowledge and Trust Metrics Pre- to Post-Video. Participants were enrolled in three cohorts: metastatic breast cancer (**A, D, G**), lung cancer (**B**,**E**,**H**), and other cancer (**C**,**F, I**). Participants completed survey assessments pre-video viewing (T1) and post-video viewing (T2) of the 3-4 minute tumor genomic testing educational video intervention. Survey assessments included: 10-question video message-specific knowledge (VMSK; **A-C**), 10-question general genomic knowledge/understanding (GKU; **D-F**), and 11-question trust in physician/provider (TIPP; **G-I**). Paired scores for each participant are presented with mean and Wilcoxon signed rank.

### Change in Individual Video Message-Specific Knowledge Questions

To further understand what specific domains of knowledge related to the message changed over time, we evaluate the proportion of patients correctly answering each of the 10 true/false questions in the video message-specific knowledge survey at T1 versus T2 (**Table 2**). The proportion correct significantly increased (McNemar’s test p<0.05) after viewing the video for five questions, including one general knowledge question (“We have genes in every cell of our bodies”), questions specifically addressing ASCO/ACMG guidelines (“Tumor tissue genomic results sometimes raise more questions that require more genetic testing”; “When my doctor has my results, they might recommend for me to see a genetics specialist”) and one question addressing cost of TGT (“The expense of TGT is not typically covered by health insurance”). The latter question addressing cost of TGT demonstrated the greatest change, increasing from 51.3% (77/15) of participants answering correctly pre-video/T1 to 80.1% (121/150) post-video/T2, an improvement of 44 individuals answering the question correctly.

**Table 2.** Change in Video Message-Specific Knowledge Responses by Individual Question.

| Video Message-specific question<br>True or False | Addresses<br>ASCO/ACMG<br>guidelines | Participants<br>correct at<br>T1 |  | Participants<br>correct at<br>T2 |  | T1:T2<br>change | p-value |
| --- | --- | --- | --- | --- | --- | --- | --- |
|  |  | # | % | # | % |  |  |
| "We have genes in every cell of our bodies." | N | 139 | 92.7 | 150 | 100 | +11 | **0.0009** |
| "TGT might help your doctor make decisions about your cancer treatment." | Y | 149 | 99.3 | 150 | 100 | +1 | 0.3173 |
| "TGT always determines what treatment a person will have." | Y | 115 | 76.7 | 130 | 86.7 | +15 | **0.0137** |
| "I must have TGT to continue with cancer treatment." | Y | 133 | 88.7 | 138 | 92.0 | +5 | 0.2253 |
| "My doctor has other tools besides TGT to use to choose treatments for me." | Y | 144 | 96.0 | 145 | 96.7 | +1 | 0.7630 |
| "Tumor tissue genomic results sometimes raise more questions that require more genetic testing." | Y | 128 | 85.3 | 145 | 96.7 | +17 | **0.0011** |
| "The information that I get from tumor tissue genomic testing could be valuable to my children and other family members." | Y | 145 | 96.7 | 149 | 99.3 | +4 | 0.1025 |
| "When my doctor has my results, they might recommend for me to see a genetics specialist." | Y | 131 | 87.3 | 147 | 98.0 | +13 | **0.0006** |
| "The expense of TGT is not typically covered by health insurance." | N | 77 | 51.3 | 121 | 80.1 | +44 | **<0.0001** |
| "If you do not have health insurance, you cannot have TGT performed." | N | 142 | 94.7 | 140 | 93.3 | -2 | 0.5930 |

ASCO: American Society of Clinical Oncology; ACMG: American College of Medical Genetics; T1: Time point 1 (pre-video); T2: Time point 2 (post-video)

### Association of Patient Characteristics with Video Message-Specific Knowledge, Genomic Knowledge/Understanding, and Trust in Physician/Provider

As an exploratory objective, the association of patient characteristics with baseline video message-specific knowledge, genomic knowledge/understanding, and Trust in Physician/Provider were assessed via Spearman correlation (**Table 3**). There was a significant association between education and baseline/T1 video message-specific knowledge (nominal p<0.0001) and baseline/T2 genomic knowledge/understanding (nominal p=0.02) as well as income and baseline/T1 video message-specific knowledge (nominal p=0.002). There were no other significant associations between other patient characteristics and video message-specific knowledge/genomic knowledge/understanding or any patient characteristics and Trust in Physician/Provider. To further evaluate these associations, we explored change in video message-specific knowledge and genomic knowledge/understanding within individual patient characteristics; no formal statistical assessment was performed for these exploratory analyses. Among individuals with educational attainment of less than a college degree (some high school, high school completion, some college or technical school; n=92) there was a greater numerical increase in video message-specific knowledge (mean 83.0 to 92.5) than among those with a college or graduate degree (mean 92.9 to 97.2) (**Supp Fig 2A**). When evaluating age, there were similar increases in video message-specific knowledge among those under age 70 at study entry (mean 87.5 to 93.9) and over age 70 at study entry (mean 85.0 to 95.5) suggesting similar knowledge receipt across the age continuum (**Supp Fig 2B**). Evaluating income above/below $75,000 per year (**Supp Fig 2C**) and race white/non-white (**Supp Fig 2D**) largely mirrored the overall population with numerical increase in video message-specific knowledge but no change in genomic knowledge/understanding.

**Table 3.** Association of Participant Characteristics with Baseline Survey Metrics.

| **Instrument** | **Demographic Parameter** | ***Nominal p-value\**** |
| --- | --- | --- |
| Video Message-Specific Knowledge at T1 | Age at entry | 0.14 |
|  | Sex | 0.36 |
|  | Education | **<0.0001** |
|  | Income | **0.002** |
|  | Marital status | 0.54 |
|  | Race | 0.16 |
| Genomic Knowledge and Understanding at T1 | Age at entry | 0.78 |
|  | Sex | 0.57 |
|  | Education | **0.02** |
|  | Income | 0.10 |
|  | Marital status | 0.88 |
|  | Race | 0.62 |
| Trust in Physician Provider at T1 | Age at entry | 0.90 |
|  | Sex | 0.55 |
|  | Education | 0.33 |
|  | Income | 0.42 |
|  | Marital status | 0.49 |
|  | Race | 0.45 |

\*Spearman correlation; T1: Time point 1 (pre-video)

### Patient Assessment of and Attituades Toward Tumor Genomic Testing

In addition to objective metrics, participants also completed 10 descriptive questions assessing their perceived knowledge and knowledge insufficiency around TGT and perceptions of TGT, and an eight question assessment of the video itself. These variables are perceived knowledge and knowledge insufficiency, which are from the Risk Information Seeking and Processing Model.^22^ Participants reported their perceived knowledge of TGT on a scale from zero to ten, with 10 meaning knowing everything that there was to know about TGT. Using the same scale, they also indicated how much they felt that they needed to know. There was a significant increase in participants’ perceived knowledge (mean T1 2.7 to T2 4.8; t-test p<0.001) while need for knowledge did not significantly increase (mean T1 5.8 to T2 6.2; t-test p=0.23) (**Supp Fig 3A**). As an attitudinal measure of participants’ perception of TGT, patients responded to the statement “My having tumor genomic testing would have…” on a scale from negative three to three, where negative three indicated “a lot more negatives than positives” and three indicated “a lot more positives than negatives”, with zero representing an equivalent number of negatives and positives (**Supp Fig 3B**). Average response value at T2 increased from 2.1-2.4, with 17% of participants having a more positive response.

To evaluate clarity of the video message being delivered and to ensure that survey respondents had indeed viewed the video (manipulation check), participants were simply asked “what was the video about?” 148 of 150 respondents correctly identified that it was about tumor genomic testing, 2 respondents chose the option “screening for cancer,” both of which were considered acceptable. A Likert scale of agreement was used to measure participant’s opinions of the video’s utility, clarity, and engagement (**Supp Fig 3C**) and most (>80%) of respondents agreed or strongly agreed that the video was helpful, easy to understand, and held their attention, with fewer but still most (60%) indicating that the graphics were helpful. Most participants (141 of 150) felt that the amount of information presented was adequate, while four participants felt the content was excessive, and five participants found the content insufficient.

## Discussion

TGT has increasingly become standard-of-care for most patients with advanced cancer and select patients with early stage cancer (including lung cancer). As TGT usage increases, there is an increasing burden on medical oncology providers to provide counseling, which is particularly challenging in community settings where access to genomic experts and genetic counseling may be more limited.^23^ Further, there is significant variability in how frequently providers educate patients prior to TGT and the content of that education.^19,24-27^ Despite consensus across international oncology and medical genetics societies around patient education prior to TGT,^3-9^ few strategies have emerged that address key guidelines while also being feasible within the flow of busy oncology clinical settings. To address this critical gap, we demonstrate that our concise, 3-4 minute animated video, now publicly released for widespread use, effectively conveys key information in an efficient timeframe to provide patient education prior to TGT.

Our results demonstrate a significant improvement in video message-specific knowledge consistent across cohorts, with >80% of participants correctly answering the 10 video message-specific knowledge questions after video viewing. As an internal control, genomic knowledge/understanding did not significantly change suggesting that knowledge gained was specifically related to the video. While individuals had a good overall baseline message-specific knowledge, the ideal would be perfect knowledge prior to TGT and significant gaps improved with video viewing. Further, most of the improvement in video message-specific knowledge score related to five questions encompassing diverse themes. It is notable that the significant increase in the video message-specific knowledge score indicates that the video is educating study participants beyond any discussion that participants had or may have had with their providers regarding TGT. These primary outcome results are further substantiated by participants’ self-report of knowledge gain and a narrowing between the knowledge participants desired to have and the knowledge participants felt they needed to be effective partners in their own care.

Videos have been recognized as an effective form of patient education decision-making tools, including specifically around genomic analyses.^28-30^ The differences between more comprehensive pre-TGT education (e.g. the Multimodality COMET eHealth Education Intervention)^31^ and concise approaches such as this video^19^ reflect a challenging balance between ensuring adequate content but also facilitating delivery within clinic flow. Shared-decision making encompasses the need for patients to discuss their concerns with family members and supporters^28,32^ and this video can be directly shared with family and friends with no need for the patient to synthesize information before sharing it with support persons. This video intervention clearly enhanced knowledge surrounding TGT as a publicly-available resource,

In prior work, we had found that patients who underwent TGT without therapy change (representing the majority of patients) lost confidence in treatment.^33^ Stronger patient-provider therapeutic alliance results in improved adherence to therapy,^34^ caregiver coping,^35^ and cancer outcomes.^36^ We did not see significant change in a ‘trust in physician/provider’ metric.^21^ We hypothesize that TIPP may not change over the narrow time frame of pre-/post-video but may become evident later in the treatment course. We plan to assess TIPP again at 60-90 days post-video. We did not specifically investigate other patient-centered outcomes as our previous prospective decision analysis study demonstrated that validated metrics of depression scale (CES-D),^37^ anxiety (BAI),^38^ and self-efficacy (CASE-cancer)^39^ did not significantly change after TGT. This was similar to the results of the ECOG-ACRIN NCI Community Oncology Research Program EAQ152 study, a randomized trial of web-based genetic education versus usual care in advanced cancer patients undergoing tumor genetic testing, that found that a web-based video intervention increased patient understanding but did not significantly reduce anxiety, depression, or cancer-specific distress.^31^ Future research should focus on defining the educational goal or PRO outcome regarding TGT which may include improving knowledge, shared decision-making, reducing anxiety/distress, increasing use of genomically directed treatments, or some combination.

Patients with lower socioeconomic status and education have less understanding of genetic/genomic testing, emphasizing the need to equitably address patient understanding prior to TGT.^34^ Less knowledge and confidence has been associated with lower rates of TGT and therefore less opportunity for TGT-directed guideline concordant care.^40^ Having a standardized video, using plain language to accommodate learners across health and genetic literacy levels, could result in more equitable enumeration of TGT benefits, limitations, possible outcomes, and risks. The exploratory analyses here demonstrated improvement in VMSK scores across demographics including income, education, and age with numerical narrowing of baseline knowledge gaps which may translate to more informed care and greater patient empowerment.

### Limitations

Our racial demographic minority subsets were not robust enough to allow for meaningful data analysis independently, therefore the 18 participants identifying as a race other than white were consolidated into one non-white category. This is not ideal and did not reveal the disparity in genomic knowledge that was seen in previous work.^32^ While a direct translation Spanish language video was available, only one Spanish-speaking participant was accrued, thus no generalizations regarding the quality or effectiveness of the translated video can be made. This study was conducted at a single tertiary academic cancer center in a large city. Because provider genomics knowledge varies by setting,^41^ we have expanded implementation of the intervention to community cancer centers. Overall, participants’ opinions of the video affirmed that length and content were sufficient, however a gap between the T1 need-for-knowledge response average and the T2 metric average suggested that patients’ may in fact desire more knowledge. We are currently collecting data on T3 (60-90 days post-TGT) which will be reported in the future to evaluate retention of knowledge.

## Conclusions

This novel TGT video intervention is an effective tool to augment the provider-patient discussion of TGT. It narrows the gap in equitable access to informed health care across several demographics: age, education, and income, but we have not demonstrated a substantial improvement in equitability across race and ethnicity. This video may be a valuable resource to facilitate awareness, enhance dialogue, extend information from the clinic to the home setting, and aid in the shared-decision making that is fundamental to patient-centered care.

## Supporting information

Suppfig1_accrual

Suppfig2_subsets

suppfig3_perceptions

## Data Availability

All data produced in the present study are available upon reasonable request to the authors.

http://www.tumor-testing.com

## Supplementary Material

### Supplementary Figure Legends

**Supplementary Figure 1. Study Accrual Features. (A)** CONSORT diagram of study enrollment. (**B**) Study accrual by month with cohort indicated. **(C)** Distribution of tumor type among 150 participants evaluable for primary endpoint; Cx=cancer. (**D)** Distribution of tumor genomic testing assay among 150 participants evaluable for primary endpoint.

**Supplementary Figure 2. Change in Knowledge Metrics Within Demographic Gropus**. Participants completed survey assessments pre-video viewing (T1) and post-video viewing (T2) of the 3-4 minute tumor genomic testing educational video intervention. Survey assessments included: 10-question video message-specific knowledge (VMSK), 10-question general genomic knowledge and understanding (GKU). Mean scores are presented for education stratified by completion of a 4-year college degree versus not (**A)**, age less than 70 years old at study entry versus greater than or equal to 70 (**B**), self-reported income less than $75,000 per year versus greater than or equal to $75,000 per year (**C**), and self-reported race non-white versus white (**D**). No statistical assessment is reported for these exploratory analyses.

**Supplementary Figure 3. Participant perceptions of knowledge, testing value, and video. (A**) Participants were asked to self-rate their knowledge and need for knowledge around tumor genomic testing (TGT) pre-video viewing (T1) and post-video viewing (T2). Box plot with mean score indicated by “x” provided, with whiskers indicating 25^th^/75^th^ percentiles and outliers indicated by dot. (**B**) Participants responded to the question, “My having tumor genomic testing would have…” and assessed the value of TGT. (**C**) Participants assessed the video quality on a 5-point Likert scale.

**Supplementary Table 1.** Change in Video Message-Specific Knowledge, Genomic Knowledge and Understanding, and Trust in Physician/Provider.

|  | Score type | T1 |  | T2 |  | Change (T2-T1) |  | P-value |
| --- | --- | --- | --- | --- | --- | --- | --- | --- |
|  |  | median | IQR | median | IQR | median | IQR |  |
| ALL | VMSK (primary) | 90 | 80, 100 | 100 | 90, 100 | 10 | 0, 10 | **<0.0001** |
|  | GKU | 80 | 80, 90 | 90 | 80, 100 | 0 | -10, 10 | 0.8905 |
|  | TIPP | 48 | 43, 52 | 48 | 43, 53 | 0 | -1, 1 | 0.5876 |
| Breast | VMSK | 90 | 80, 100 | 100 | 90, 100 | 10 | 0, 10 | **<0.0001** |
|  | GKU | 90 | 80, 100 | 90 | 80, 100 | 0 | -10, 0 | 0.8866 |
|  | TIPP | 50 | 44, 53 | 49 | 45, 54 | 0 | -1, 1 | 0.5891 |
| Lung | VMSK | 85 | 70, 90 | 90 | 90, 100 | 10 | 0, 10 | **<0.0001** |
|  | GKU | 90 | 80, 90 | 90 | 80, 100 | 0 | 0, 10 | 0.9547 |
|  | TIPP | 44.5 | 40, 48 | 44.5 | 40, 51 | 0 | -1, 1 | 0.8913 |
| Agnostic | VMSK | 90 | 80, 100 | 100 | 90, 100 | 10 | 0, 10 | **<0.0001** |
|  | GKU | 90 | 80, 100 | 90 | 80, 100 | 0 | -10, 10 | 0.8173 |
|  | TIPP | 50 | 43, 53 | 50 | 44, 53 | 0 | -1, 2 | 0.1402 |

VMSK: Video message-specific knowledge; GKU: General genomic knowledge and understanding; TIPP: Trust in Physician/Provider

