## Supplementary figures and images for "A Video Intervention to Improve Patient Understanding of Tumor Genomic Testing in Patients with Cancer"

### Suppfig1_accrual

**Supplementary Figure 1.**

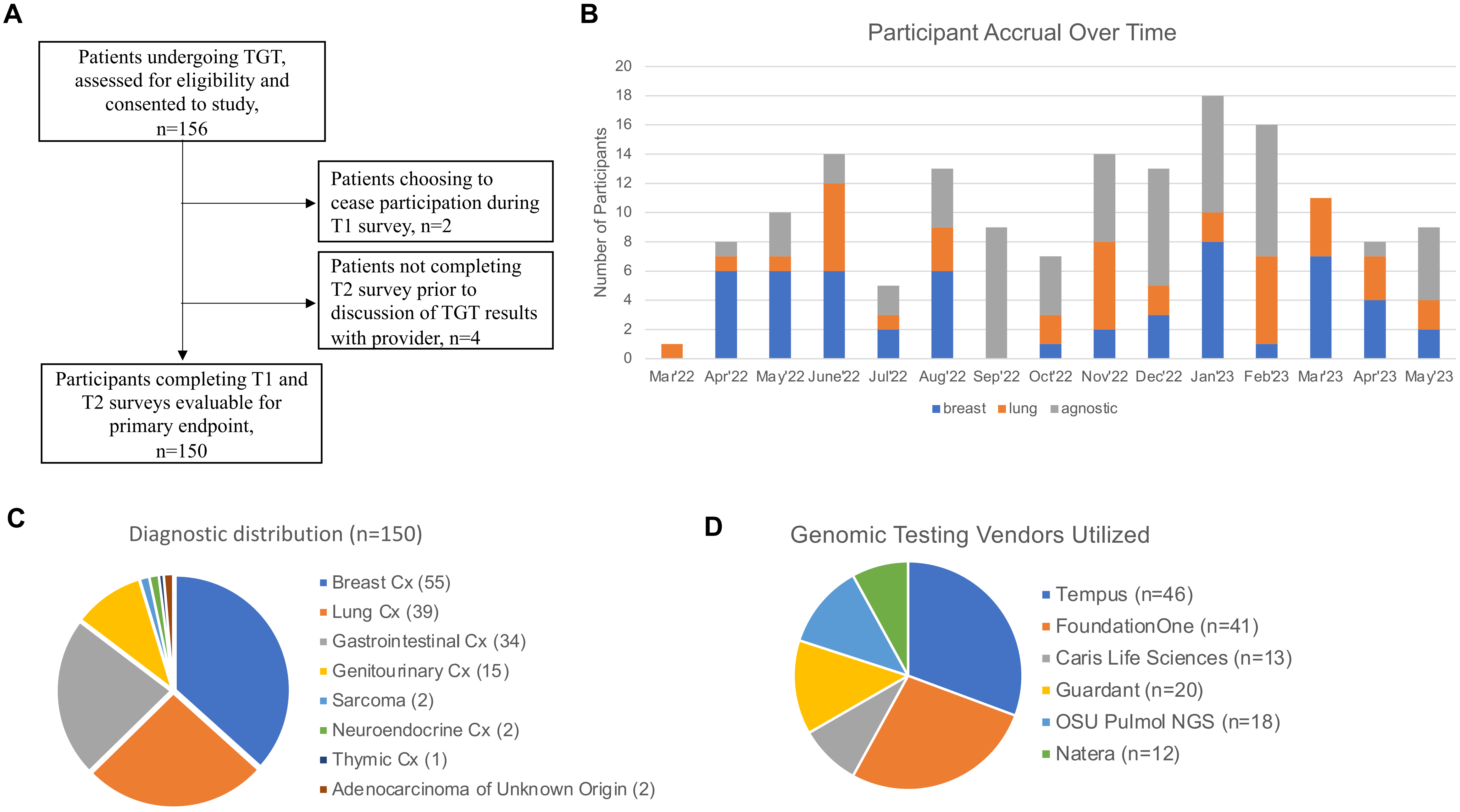

### Suppfig2_subsets

#### Supplementary Figure 2.

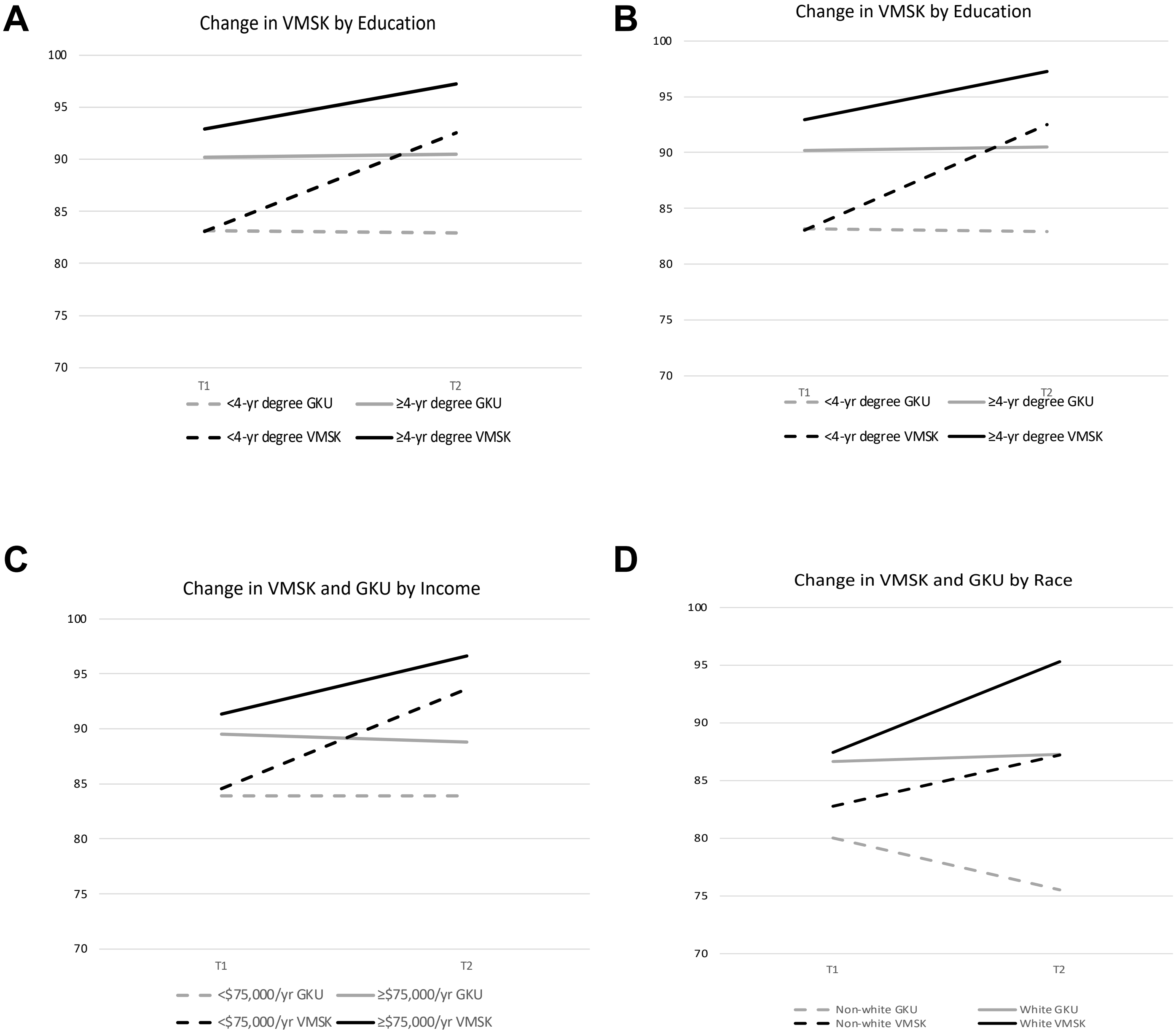

### suppfig3_perceptions

#### Supplementary Figure 3.

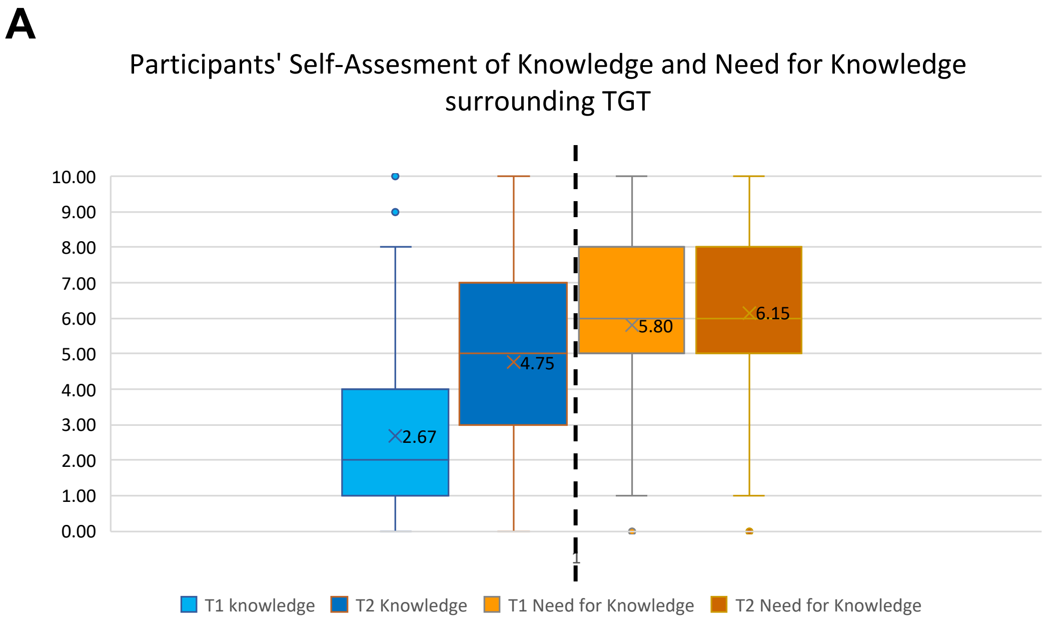


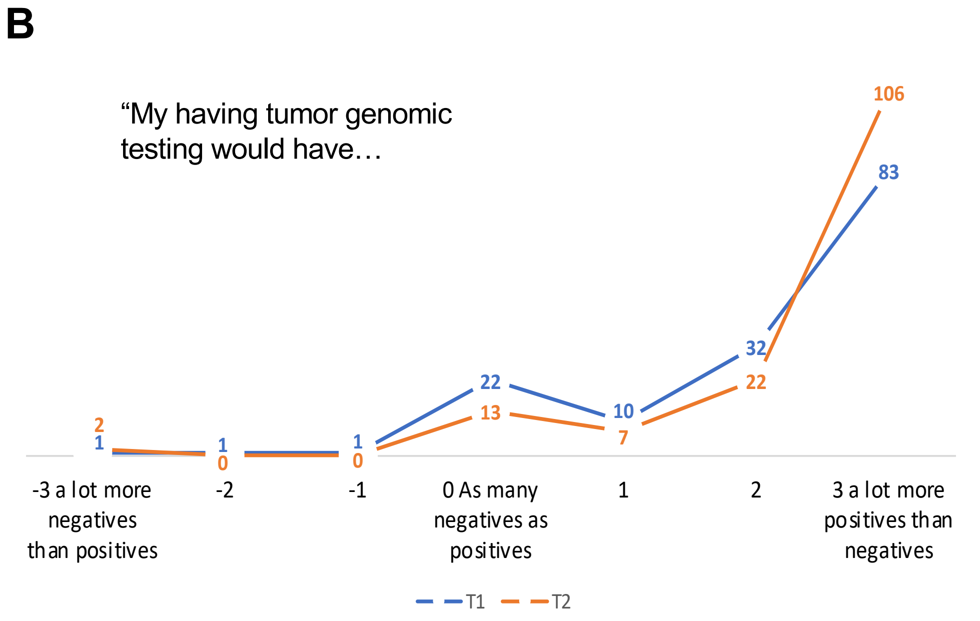


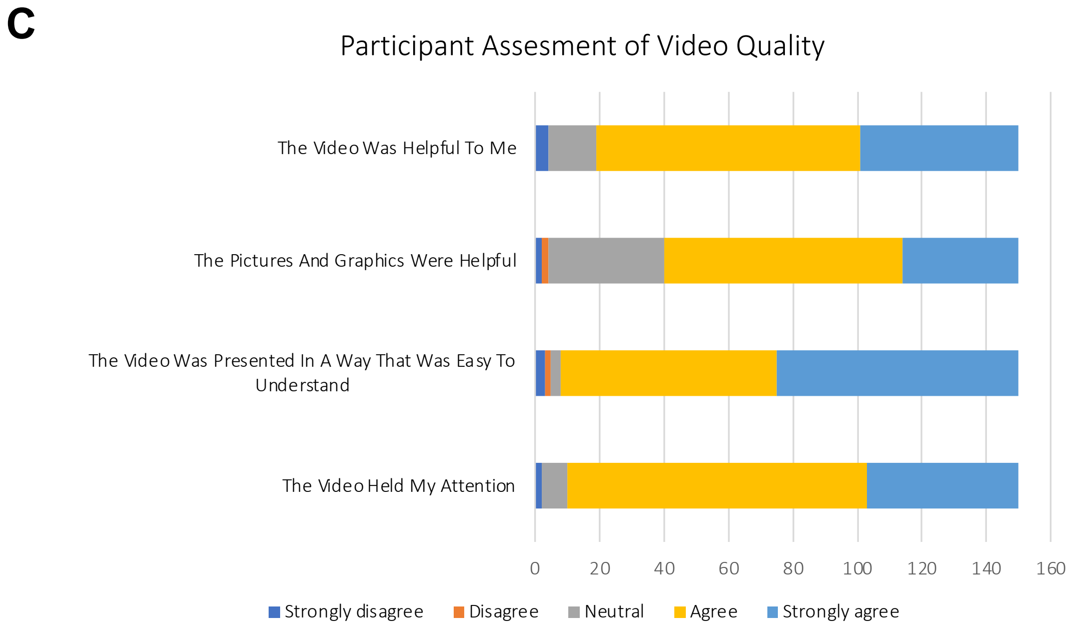
